# Active or latent tuberculosis increases susceptibility to COVID-19 and disease severity

**DOI:** 10.1101/2020.03.10.20033795

**Authors:** Yu Chen, Yaguo Wang, Joy Fleming, Yanhong Yu, Ye Gu, Chang Liu, Lichao Fan, Xiaodan Wang, Moxin Cheng, Lijun Bi, Yongyu Liu

## Abstract

**Importance:** Risk factors associated with COVID-19, the viral pneumonia originating in Wuhan, China, in Dec 2019, require clarification so that medical resources can be prioritized for those at highest risk of severe COVID-19 complications. Infection with *M. tuberculosis* (MTB), the pathogen that causes TB and latently infects ∼25% of the global population, may be a risk factor for SARS-CoV-2 infection and severe COVID-19 pneumonia.

**Objective:** To determine if latent or active TB increase susceptibility to SARS-COV-19 infection and disease severity, and lead to more rapid development of COVID-19 pneumonia.

**Design:** An observational case-control study of 36 confirmed COVID-19 cases from Shenyang, China, conducted in Feb 2020. Final date of follow-up: Feb 29, 2020. Cases were grouped according to COVID-19 pneumonia severity (mild/moderate, severe/critical), and MTB infection status compared. Comparisons were made with MTB infection data from another case-control study on bacterial/viral pneumonia at Shenyang Chest Hospital.

**Setting:** Multi-center study involving three primary care hospitals in Shenyang, China.

**Participants:** 86 suspected COVID-19 cases from participating primary-care hospitals in Shenyang. All 36 SARS-CoV-2 +ve cases (based on RT-PCR assay) were included. Disease severity was assessed using the Diagnostic and Treatment Guidelines of the National Health Commission of China (v6). Mean age, 47 years (range: 25-79), gender ratio, 1:1.

**Exposures:** Confirmed COVID-19 pneumonia. Interferon-gamma Release Assays (IGRA) were performed using peripheral blood to determine MTB infection.

**Main Outcome and Measures:** Epidemiological, demographic, clinical, radiological, and laboratory data were collected. Comparison of MTB infection status between patients with mild/moderate and severe/critical COVID-19 pneumonia.

**Results:** Mean age of 36 COVID-19 patients: 47 (range: 25-79); M/F: 18/18; Wuhan/Hubei connection: 42%. Mild/moderate cases: 27 (75%); severe/critical: 9 (25%). MTB infection (IGRA+ve): 13 cases (36.11%), including 7 of 9 severe/critical cases. MTB infection rate: higher in COVID-19 (36.11%) than bacterial pneumonia (20%; p=0.0047) and viral pneumonia patients (16.13%; p=0.024). MTB infection more common than other co-morbidities (36.11% vs diabetes: 25%; hypertension: 22.2%; coronary heart disease: 8.33%; COPD: 5.56%). MTB co-infection linked with disease severity (severe/critical 78% vs mild/moderate cases 22%; p=0.0049), and rate of disease progression: infection to development of symptoms (MTB+SARS-CoV-2: 6.5±4.2 days vs SARS-COV-2: 8.9±5.2 days; p=0.073); from symptom development to diagnosed as severe (MTB+SARS-CoV-2: 3.4±2.0 days vs SARS-COV-2: 7.5±0.5 days; p=0.075).

**Conclusions and Relevance:** MTB infection likely increases susceptibility to SARS-CoV-2, and increases COVID-19 severity, but this requires validation in a larger study. MTB infection status of COVID-19 patients should be checked routinely at hospital admission.

**Key Points:** *Question:* Is latent or active tuberculosis (TB) a risk factor for SARS-CoV-19 infection and progression to severe COVID-19 pneumonia?

*Findings:* In this observational case-control study of 36 COVID-19 cases from Shenyang, China, we found tuberculosis history (both of active TB and latent TB) to be an important risk factor for SARS-CoV-2 infection. Patients with active or latent TB were more susceptible to SARS-CoV-2, and COVID-19 symptom development and progression were more rapid and severe.

*Meaning:* Tuberculosis status should be assessed carefully at patient admission and management and therapeutic strategies adjusted accordingly to prevent rapid development of severe COVID-19 complications.

## Introduction

COVID-19, the viral pneumonia that emerged in Wuhan, China, in December 2019, has spread throughout China and to 85 other countries. To date (Mar 5, 2020), there have been 95,333 confirmed cases worldwide and 3282 deaths^1^. COVID-19 poses a major threat to global health and was declared a ‘public health emergency of international concern’ by the WHO on 30 Jan 2020^2^. While most SARS-CoV-2 infections in the general population result in only mild symptoms, individuals with underlying comorbidities, particularly the elderly, are more vulnerable to SARS-CoV-2 infection and require additional care^3-5^. It is important to gain a clear picture of risk factors associated with this new viral respiratory infection in order to allocate appropriate medical resources to prevent the development of severe or critical forms of COVID-19 in those considered to be at higher risk. Here, we consider the possibility that infection with *Mycobacterium tuberculosis*, the pathogen that causes tuberculosis (TB), the top cause of death due to an infectious disease, may predispose to SARS-CoV infection and the more rapid development of symptoms and severe COVID-19 pneumonia.

## Methods

### Data collection

86 people with COVID-19 symptoms presented at hospitals in Shenyang, China, between Jan 26, 2020 and Feb 15, 2020 and were tested for SARS-CoV-2 infection. COVID-19 infection was confirmed in 36 cases (Shenyang Sixth People’s Hospital [32 cases], Shenyang Fourth People’s Hospital [4 cases] and Shenyang Chest Hospital [1 case]) and patients were hospitalized and enrolled in the study. Date of admission, gender, age, laboratory test results, and chest radiography results were extracted from patient admission records. The number of days from presumed contact with a suspected/confirmed COVID-19 patient to symptom development, the date on which SARS-CoV-2 infection was confirmed, and disease severity were also recorded.

Data on MTB infection status were compared with retrospective data obtained from a case-series study of 115 bacterial and 62 other viral pneumonia patients conducted in the Pulmonology Department of Shenyang Chest Hospital (unpublished data). All patients diagnosed with bacterial or viral pneumonia from Dec 20, 2019 to Feb 20, 2020 (cases in which SARS-CoV-2 infection/COVID-19 disease was ruled out according to criteria in the Diagnostic and Treatment Guidelines of the National Health Commission of China (v5) ^9^) were included in the study. Demographic data are shown in Supplementary Table 1.

These studies were approved by the Ethics Committee of Shenyang Chest Hospital, and informed written consent was obtained from each of the study participants.

### SARS-CoV-2 testing

Throat swabs, blood and sputum were collected from suspected COVID-19 patients and sent for SARS-CoV-2 testing using a real-time reverse-transcription polymerase chain reaction assay^6,7^at the Shenyang Disease Control Center.

### IFN-γ IGRA assays

IFN-γ IGRA assays were performed using X.DOT-TB kits (TB Healthcare, Foshan, China) according to the manufacturer’s instructions^8^.

### Statistical analysis

Analyses were performed using R (version 3.6.1). The normality of data distributions was assessed using the Kolmogorov-Smirnov test. Normally-distributed data are presented as means (SD), while non-normally distributed data are presented as medians (IQR), and categorical variables as frequencies (%). Differences between groups were analysed by Fisher’s exact test or Pearson’s chi-squared test (for categorical data), and Students’ *t*-tests or Mann-Whitney tests (for continuous data).

## Results

Among the 36 confirmed COVID-19 patients from Shenyang, Liaoning province, China, included in this study, the most common symptoms at presentation were cough (61.11%), dyspnea (33.33%), fever (27.78%), lymphopenia (27.78%) and fatigue (25%) (Table 1). The mean age of patients was 47 years (range: 25-79), and 18 were men. Fifteen cases had a connection with Wuhan/Hubei province or neighboring cities, while 21 had become infected via local transmission (3), or from friends/family members in Shenyang already diagnosed with COVID-19.

**Table 1.** Comorbidities present in COVID-19 patients

| Comorbidities | All patients<br>(n=36) | Mild/Moderate<br>(n=27) | Severe/Critical<br>(n=9) | *p* value*<br>(Fisher's exact<br>test) |
| --- | --- | --- | --- | --- |
| Tuberculosis (TB) | .. | .. | .. | 0.0049 |
| ELISPOT positive | 13 (36.11%) | 6 (22.22%) | 7 (77.78%) | .. |
| old TB calcifications | 3 | 2 | 1 | .. |
| TB history | 5 | 2 | 3 | .. |
| active TB | 3 | 0 | 3 | .. |
| LTBI | 2 | 2 | 0 | .. |
| ELISPOT negative | 23 (63.89%) | 21 (77.78%) | 2 (22.22%) | .. |
| Chronic obstructive<br>pulmonary disease<br>(COPD) | .. | .. | .. | 0.0571 |
| positive | 2 (5.56%) | 0 (0) | 2 (22.22%) | .. |
| negative | 34 (94.44%) | 27 (100%) | 7 (77.78%) | .. |
| Diabetes | .. | .. | .. | 0.738 |
| positive | 9 (25.00%) | 7 (25.93%) | 2 (22.22%) | .. |
| negative | 27 (75%) | 20 (74.07%) | 7 (77.78%) | .. |
| Hypertension | .. | .. | .. | 0.6478 |
| positive | 8 (22.22%) | 7 (25.93%) | 1 (11.11%) | .. |
| negative | 28 (77.78%) | 20 (74.07%) | 8 (88.89%) | .. |
| Coronary heart disease | .. | .. | .. | 1 |
| positive | 3 (8.33%) | 2 (7.40%) | 1 (11.11%) | .. |
| negative | 33 (91.67%) | 25 (92.60%) | 8 (88.89%) | .. |
| Cerebrovascular diseases | 0 | 0 | 0 | .. |
| Hepatitis B infection | .. | .. | .. | 0.25 |
| positive | 1 (2.78%) | 1 (3.7%) | 0 (0%) | .. |
| negative | 35 (97.22%) | 26 (96.3%) | 9 (100%) | .. |
| Cancer | 0 | 0 | 0 | .. |
| Chronic renal diseases | 0 | 0 | 0 | .. |
| Immunodeficiency | .. | .. | .. | 0.25 |
| positive | 1 (2.78%) | 1 (3.70%) | 0 (0%) | .. |
| negative | 35 (97.22%) | 26 (92.30%) | 9 (100%) | .. |

Data presented: n (%). \*Significance of differences between the Mild/Moderate and Severe/Critical groups.

To evaluate the implications of MTB infection history on the incidence and progression of COVID-19 disease, we addressed three questions. Does MTB co-infection: 1. increase susceptibility to SARS-COV-19 infection; 2. increase disease severity; and 3. lead to more rapid disease progression? We first assigned cases into two groups based on the severity of their COVID-19 pneumonia according to the Diagnostic and Treatment Guidelines of the National Health Commission of China (v6) ^9^. Twenty-seven cases were mild/moderate (75%, Group 1) and nine cases were severe/critical (25%, Group 2) (Table 2). We then determined the MTB infection status of each patient by examining their clinical history and by performing X.DOT-TB IGRA assays (Table 1). Thirteen of the 36 COVID-19 cases (36.11%) were IGRA+ve, of which 3 had active TB (1 MDR-TB), and 5 were recovered TB patients. Old TB calcifications were found on chest scans of 3 patients who had not previously been diagnosed with TB, and 2 patients had latent TB (LTBI). The percentage of IGRA+ve COVID-19 cases is approximately double the estimate of the percentage of IGRA+ve individuals in the general population (15-18% in rural China^10^), suggesting that MTB infection likely is an important risk factor for susceptibility to SARS-COV-2 infection. To determine if MTB infection is a risk factor specific for COVID-19 pneumonia, or for pneumonia in general, we compared the MTB infection rate within these COVID-19 cases with data from a case-series comprised of 115 bacterial pneumonia and 62 viral pneumonia patients obtained from another study conducted in Shenyang (unpublished) (Figure 1A); MTB infection rates were considerably higher among COVID-19 patients than among bacterial pneumonia patients (36.11% vs 20%; *p* = 0.047 Figure 1A) and viral pneumonia patients (36.11% vs 16.13%; *p* = 0.024), suggesting that MTB infection status is a specific risk factor for SARS-COV-2 infection rather than for pneumonia in general. The rate of MTB infection in the bacterial and other viral pneumonia patients was not significantly different and was approximately the same as that in the general population^10^. The percentage of patients with MTB infection was also considerably higher than the percentage of patients with other co-morbidities (diabetes: 25%; hypertension: 22.2%; coronary heart disease: 8.33%; COPD: 5.56%; Table 1), suggesting MTB infection represents a higher risk for SARS-CoV-2 infection.

**Table 2.** Clinical characteristics and laboratory results for 36 COVID-19 cases

| Clinical Characteristics | All cases<br>(n = 36) | Mild/Moderate<br>(n = 27) | Severe/Critical<br>(n = 9) | *p*-value* |
| --- | --- | --- | --- | --- |
| Age (years) | 47±14 | 48±15 | 56±11 | 0.8856 |
| Gender (M/F) | 18/18 | 16/11 | 7/2 | 0.4377 |
| Fever | 23 (63.89%) | 16 (59.26%) | 7 (77.78%) | 0.4377 |
| Highest temperature during hospital admission | 37.7±0.99 | 37.6±1.03 | 37.9±0.89 | 0.5091 |
| Conjunctival congestion | 3 (8.33%) | 1 (3.70%) | 2 (22.22%) | 0.1866 |
| Nasal congestion | 5 (13.89%) | 5 (18.52%) | 0 | 0.4908 |
| Headache | 2 (5.56%) | 2 (7.41%) | 0 | 1 |
| Cough | 22 (61.11%) | 18 (66.67%) | 4 (44.44%) | 0.7797 |
| Sore throat | 7 (1.44%) | 5 (18.52%) | 2 (22.22%) | 0.7832 |
| Sputum production | 8 (22.22%) | 6 (22.22%) | 2 (22.22%) | 0.6517 |
| Fatigue | 9 (25%) | 5 (18.52%) | 4 (44.44%) | 0.4593 |
| Hemoptysis | 2 (5.56%) | 0 | 2 (22.22%) | 0.0782 |
| Dyspnea | 12 (33.33%) | 4 (14.81%) | 8 (88.89%) | 0.0235 |
| Nausea or vomiting | 5 (13.89%) | 3 (11.11%) | 2 (22.22%) | 0.8644 |
| Diarrhea | 7 (19.44%) | 5 (18.52%) | 2 (22.22%) | 0.7832 |
| Myalgia or arthralgia | 6 (16.67%) | 3 (11.11%) | 3 (33.33%) | 0.4431 |
| Chills | 5 (13.89%) | 4 (14.81%) | 1 (11.11%) | 0.7752 |
| Laboratory Tests | All cases<br>(n = 36) | Mild/Moderate<br>(n = 27) | Severe/Critical<br>(n = 9) | *p*-value* |
| Blood leukocyte count<br>(3.5-9.5×10<sup>9</sup>/L) | 6.384±3.548 | 5.928±2.602 | 8.433±5.671 | 0.0831 |
| Lymphocyte count<br>(1.1-3.2×10<sup>9</sup>/L) | 1.657±1.115 | 1.861±1.037 | 0.738±0.477 | 0.0164 |
| Lymphopenia | 10 (27.78%) | 5 (18.52%) | 5 (55.56%) | 0.0794 |
| Platelet count<br>(125-350×10<sup>9</sup>/L) | 220.75±107.077 | 216.519±110.59<br>7 | 233.444±92.93<br>7 | 0.6875 |
| Haemoglobin level<br>(115-150g/L) | 140(20) | 139.6154(19) | 140(19) | 0.6519 |
| C-reactive protein level<br>(0-4mg/L) | 5.36(27.14) | 4.47(10.37) | 35.88(95.59) | 0.002 |
| Procalcitonin level<br>(0.02-0.05ng/mL) | 0.04(0.0125) | 0.04(0.01) | 0.05(0.05) | 0.037 |
| Lactose dehydrogenase<br>(120-250U/L) | 466.429±160.61<br>3 | 455.846±143.87<br>0 | 497±200.486 | 0.4638 |
| Aspartate aminotransferase<br>(13-35U/L) | 26.743±15.483 | 23.692±12.562 | 35.556±19.475 | 0.0395 |
| Alanine aminotransferase<br>(7-40U/L) | 32.657±22.357 | 26.385±17.613 | 50.778±24.432 | 0.0027 |
| Total bilirubin<br>(3.42-20.52μmol/L) | 19.906±8.892 | 19.462±7.368 | 21.834±12.167 | 0.438 |
| Creatinine kinase (40-200U/L) | 40(27) | 39.5(23) | 59(86) | 0.104 |
| Creatinine (41-73μmol/L) | 51.706±20.446 | 51.038±15.334 | 54.6±29.976 | 0.5495 |
| Sodium (137-147mmol/L) | 132.771±22.371 | 133.154±25.374 | 131.667±3.082 | 0.2549 |
| Potassium (3.5-5.3mmol/L) | 4.312±1.457 | 4.327±0.972 | 4.25±2.145 | 0.7467 |
| Chloride (99-110mmol/L) | 97.8±16.846 | 97.846±18.775 | 97.66±6.595 | 0.9163 |

Data: mean ± SD or n (%). \*Difference between mild/moderate and severe/critical groups.

**Figure 1.**
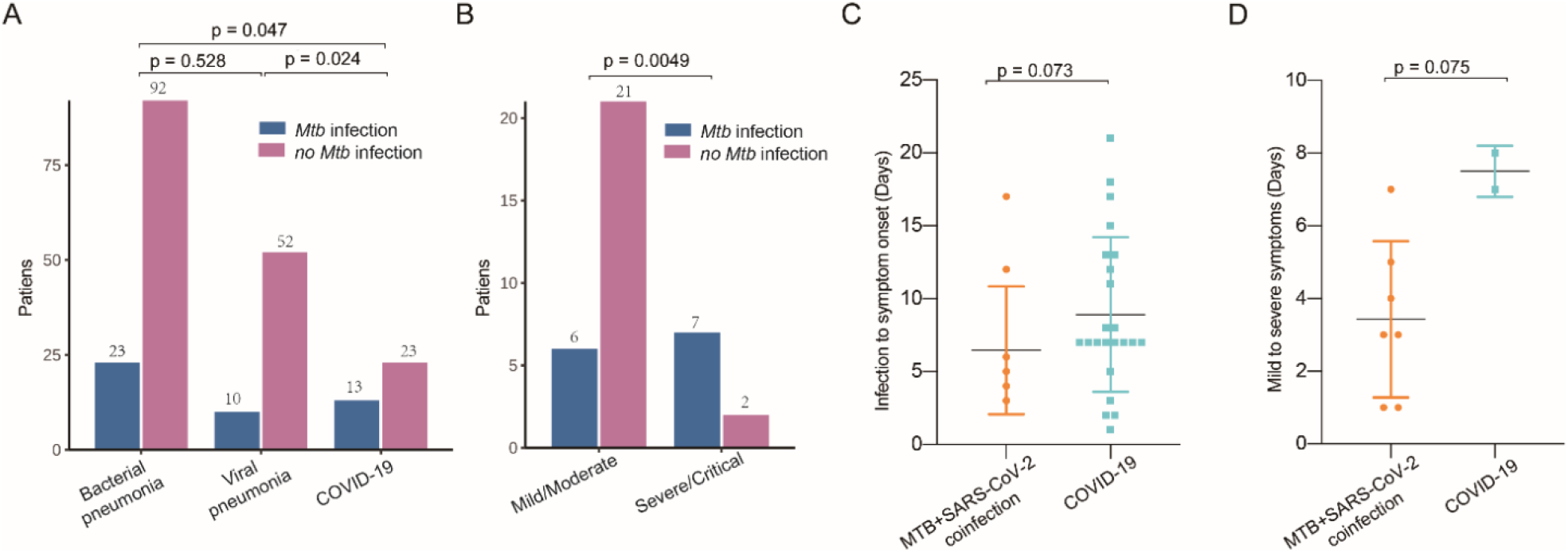
TB history affects susceptibility to SARS-CoV-2 and can increase disease severity. 36 COVID-19 patients, 115 bacterial pneumonia patients and 62 viral pneumonia patients were tested for MTB infection using IFN-γ release ELISPOT assays. IFN-γ release indicated MTB infection. **A**. MTB infection is more common among COVID-19 patients than in those with bacterial or other viral pneumonias. **B**. Patients with MTB infection developed more severe COVID-19 symptoms. **C**. COVID-19 symptoms develop more rapidly in patients with MTB coinfection. **B**. COVID-19 patients with MTB infection develop severe complications more rapidly. Comparisons between groups were made using Fisher’s exact test (n<40; p≤0.05), or the Mann-Whitney test (n>40; p≤0.05).

With respect to disease severity, the percentage of severe/critical cases with MTB co-infection was significantly higher than that in the mild/moderate group (78% vs 22%; p = 0.0049; Figure 1B). Seven of the 9 severe/critical cases were IGRA+ve indicating MTB co-infection, and of these, 4 of the 5 cases that developed ARDS were IGRA+ve. By comparison, 21 of 27 cases (78%) in Group 1 (mild/moderate cases) were IGRA-ve and had no history of TB disease. This data infers that coinfection of SARS-CoV-2 with MTB tends to lead to the development of more severe or critical COVID-19 symptoms.

Our data also show that MTB infection is associated with more rapid development of symptoms (Figure 1C-D); MTB coinfected COVID-19 cases developed symptoms on average 3.3 days earlier than their non-MTB-infected counterparts. Development of critical symptoms is reported to occur on average 9 days after initial symptom onset^4^. The 7 MTB-infected severe/critical COVID-19 cases here, however, were categorized as severe/critical 3.4 days after initial symptom development. While the number of cases here is not sufficient to draw a solid conclusion, it would appear that COVID-19 disease progresses more rapidly in the presence of MTB coinfection.

## Discussion

This observational study of the relationship between MTB infection and COVID-19 pneumonia suggests that individuals with latent or active TB may be more susceptible to SARS-CoV-2 infection, and that COVID-19 disease progression may be more rapid and severe. Given that TB causes more deaths than any other infectious disease (1.45 million deaths and 10 million new cases in 2018^11^), and that global rates of LTBI are estimated to be as high as 25%^11^, these findings are a cautionary reminder to clinicians that MTB infection status should be considered when treating COVID-19 patients in order to prevent rapid deterioration in patient health.

Our data suggest that MTB infection could be a more important risk factor than the comorbidities commonly reported in epidemiological studies such as diabetes and hypertension^12^. COVID-19 and TB are both respiratory diseases. It is perhaps not surprising that chronic respiratory diseases such as Chronic Obstructive Pulmonary Disease (COPD), and indeed active TB, could predispose susceptibility to SARS-CoV-2. Statistics from other reported studies, however, suggest that the frequency of these co-morbidities is much lower than that of MTB infection in this study (COPD: 1% of 1099 COVID-19 cases^12^, chronic respiratory disease: 2.4% of 72,314 COVID-19 cases^13^). The strength of this finding, however, is limited by the relatively low number of cases involved in this study and requires further validation.

Based on our findings, we make the following recommendations for the management and treatment of patients with a history of MTB infection (LTBI or active TB) and possible SARS-CoV-2 coinfection. First, the medical and wider community should be informed that latent/active TB is a risk factor for SARS-CoV-2 infection, and those at risk should be encouraged to pay special attention to preventive measures. Ideally, this vulnerable group should be monitored regularly at community-based medical centers in situations when resources are available. Second, the MTB infection status (history and IGRA response) of suspected/confirmed COVID-19 cases should be checked routinely on admission to ensure coinfected cases are placed in appropriate isolation wards if active TB is detected. Third, special medical resources (such as ECMO therapy) should be arranged in advance for coinfected patients in view of the increased likelihood of rapid development of severe or critical symptoms. Fourth, therapeutic approaches for cases known to be coinfected both SARS-CoV-2 and MTB should bear their MTB infection history in mind; for example, treatments involving immunosuppressive agents should be reconsidered, as they hold the potential to reactivate latent TB infections.

This is the first study to date to consider MTB infection as a comorbidity for COVID-19. Going forward, it will be important to validate the relationship uncovered here among these 36 COVID-19 cases in a larger study.

## Data Availability

All data referred to in the manuscript and provided in the manuscript files.

## Acknowledgments

We would like to thank all healthcare professionals involved in the treatment and management of the COVID-19 patients in this study for their dedication and commitment. We would like to acknowledge Dr Juanjuan Zhang, MD (Shenyang Fourth People’s Hospital) for coordinating the four COVID-19 cases in the cohort from Shenyang Fourth People’s Hospital. We also thank BC-BIO Biotechnology (Guangdong, China) for help with data analysis.

## Funding/Support

Funding for this work was from the Shenyang Major Science and Technology Innovation R&D Program (JY2020-9-018 to Y. Chen), and the National Science and Technology Major Projects (2018ZX10731301-003, 2018ZX10731301-009 and 2018ZX10731301-010 to L. Bi).

## Role of the Funder/Sponsor

The funders had no role in the design and conduct of the study; collection, management, analysis, and interpretation of the data; preparation, review, or approval of the manuscript; and decision to submit the manuscript for publication.

## Conflict of Interest Disclosures

The authors declare no conflicts of interest.

## Author Contributions

Y. Chen, Y. Wang, J. Fleming, L. Bi and Y. Liu had full access to all of the data in the study and take responsibility for the integrity of the data and the accuracy of the data analysis. Y. Chen, Y. Wang and J. Fleming contributed equally and share first authorship.

*Concept and design*: Y. Chen, Y. Wang, J. Fleming, L. Bi and Y. Liu

*Acquisition, analysis, or interpretation of data*: Y. Chen, Y. Wang, J. Fleming, Y. Yu, C. Liu, L. Fan, X. Wang, M. Cheng

*Drafting of the manuscript*: J. Fleming, Y. Wang

*Critical revision of the manuscript for important intellectual content*: Y. Chen, Y. Wang, J. Fleming, L. Bi and Y. Liu

*Statistical analysis*: Y. Wang

*Obtained funding*: Y. Chen, L. Bi

*Administrative, technical, or material support*: Y. Chen, Y. Yu, C. Liu, L. Fan, X. Wang, M. Cheng

*Supervision*: Y. Gu, L. Bi, Y. Liu

## Supplementary Information

**Supplementary Table 1.**
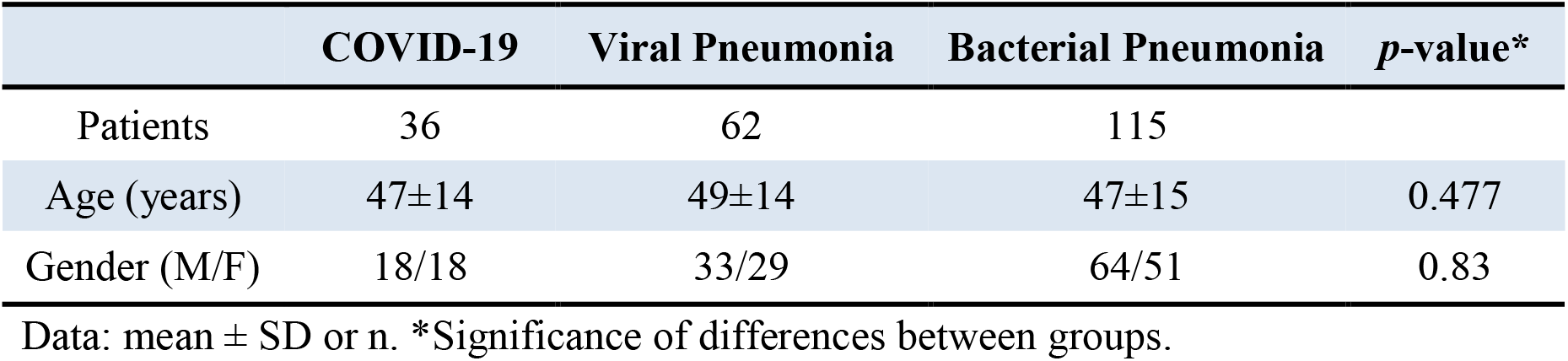
Demographic characteristics of COVID-19 and bacterial or other viral pneumonia patients

|  | COVID-19 | Viral Pneumonia | Bacterial Pneumonia | *p*-value* |
| --- | --- | --- | --- | --- |
| Patients | 36 | 62 | 115 |  |
| Age (years) | 47±14 | 49±14 | 47±15 | 0.477 |
| Gender (M/F) | 18/18 | 33/29 | 64/51 | 0.83 |

Data: mean ± SD or n. \*Significance of differences between groups.

## Notes

### Competing Interest Statement

The authors have declared no competing interest.

